# Early Detection of Rare Disease Using Hierarchical Set-to-Sequence Modeling of Structured Electronic Health Records

**DOI:** 10.64898/2026.05.04.26352393

**Authors:** Yingbo Ma, Lokesh K. Chinthala, Akram Mohammed, Robert L. Davis, Vincenza Colonna

## Abstract

Rare diseases are characterized by heterogeneous, weak, and sparse phenotypic signals that emerge gradually across longitudinal clinical visits, making early detection a persistent challenge. In this study, we propose a hierarchical set-to-sequence (HSS) framework for prospective rare disease detection using structured EHR data. HSS decomposes the problem into two levels: (1) intra-visit encoding via Multi-Query Attention (MQA), which treats heterogeneous clinical events within a single clinical visit as an unordered set to generate unified visit-level representations, and (2) inter-visit temporal modeling with transformer encoders conditioned on patient visit age and inter-visit time gaps to capture the disease progression and the irregular intervals between clinical visits. We construct a real-world cohort of 40,223 patients comprising 708,422 visits from a single academic medical center (2005–2025), with 3,032 rare disease cases identified by curated rule-based phenotyping including severe neuro-developmental, congenital, or genetic conditions. We formulate the task as multi-horizon prospective binary classification with five prediction horizons of 7, 30, 90, 180, and 365 days prior to first diagnosis. Experimental results show that the proposed HSS model consistently outperforms linear logistic regression, tree-based XGBoost, and Transformer-based baselines at every prediction horizon, ranging from AUROC = 0.893 and AUPRC = 0.601 at 7 days with 5.17% prevalence to AUROC = 0.829 and AUPRC = 0.228 at 365 days with at 3.98% prevalence. Notably, the performance gap between HSS and the strongest competing baseline is largest at the 365 days horizon, indicating stronger advantages for long-horizon prediction where phenotypic signals for rare diseases are weak and sparse. Additional analyses further clarify the contribution of the hierarchical components and confirm the importance of hierarchical modeling. This work contributes to the ongoing development of AI methodologies tailored to rare diseases by introducing a hierarchical framework for early detection using structured longitudinal clinical data.

## I. Introduction

Rare diseases collectively affect an estimated 300-400 million individuals worldwide [1]. Many individuals with rare diseases remain undiagnosed for years due to nonspecific, heterogeneous symptoms that evolve gradually across longitudinal clinical encounters and specialties, and the average time from symptom onset to confirmed rare disease diagnosis typically ranges from 4 to 7 years [2]. Consequently, patients often experience prolonged diagnostic delays, often causing chronic illness and premature death with substantial clinical, psychological, and economic burdens [3]. Therefore, it is important to detect high-risk patients early and refer for further genetic testing, such as whole-genome sequencing and gene panels [4].

Electronic health records (EHRs) provide unique perspectives into the early detection of rare diseases [5], [6]. Unlike traditional medical imaging or genomic tests, which are typically ordered after suspicion has emerged, EHR data are generated continuously during routine care. Over time, EHR data record rich information of patients’ health trajectories including heterogeneous phenotypic changes across diagnoses, laboratory results, procedures, medications, and specialties. If effectively modeled, the evolving patterns in EHR data may expose early signals of underlying rare disease and enable risk identification well before traditional diagnostic pathways are triggered [7].

Despite the potential of prospective rare disease detection using EHR data, relatively few studies have investigated this setting, and much of the existing literature focuses on retrospective classification where diagnostic signals may be already prominent [5], [6], [8], [9]. In contrast, prospective detection is inherently more challenging as early signs of rare diseases are often subtle and widely dispersed across clinical visits and specialties, with the multimodal, high-dimensional, and irregularly sampled nature of EHR data further compounding this difficulty [10], [11].

To address this gap, we propose a Hierarchical Set-to-Sequence (HSS) framework for prospective detection of rare diseases using longitudinal EHR data. This framework explicitly models the hierarchical structure of EHR by disentangling intra-visit heterogeneity from inter-visit temporal progression. Within each visit, clinical events are modeled as unordered sets to capture coordinated phenotype patterns from multimodal data, as the fine-grained intra-visit ordering of diagnoses, medications, or laboratory entries within a single encounter provides limited information for modeling the long-term progression of rare diseases [12], [13]. In contrast, across visits are modeled as temporal sequences to capture longitudinal dependencies and progressive phenotype accumulation over time. By leveraging a hierarchical set-to-sequence architecture, HSS is designed to capture the temporal accumulation of weak evidence in rare disease trajectories that precede diagnosis. We evaluated our approach under a rigorous multi-horizon prospective design, formulating rare disease detection as a binary classification task across five prediction horizons (7-, 30-, 90-, 180-, and 365-day prior to first diagnosis). Experiments are conducted on a large real-world EHR cohort comprising 40,223 patients and over 708,000 visits spanning 20 years, with rare disease cases identified through curated rule-based phenotyping. Experimental results across all prediction horizons show consistent improvements in AUROC and AUPRC over strong baselines.

## II. Related Work

### A. Rare Disease Detection Using EHR

EHR data have increasingly been used to support rare disease identification and phenotyping. Prior work has developed curated rule-based phenotyping algorithms to identify rare disease cohorts within EHR data. These approaches typically combine expert-defined diagnostic code sets and temporal criteria [5], [14]. While such rule-based strategies can achieve high precision for well-characterized conditions, they often require substantial domain expertise and are labor-intensive. In parallel, advances in genomic medicine have led to phenotype-driven gene prioritization methods, where Human Phenotype Ontology (HPO) terms are often combined with sequencing data to rank candidate causal genes [15], [16]. However, such methods typically assume that a relatively complete and curated phenotype profile is already available and often after specialist evaluation. More recently, machine learning approaches have incorporated clinical text to detect rare disease indicators from unstructured narrative data [17], [18]. These approaches can reveal nuanced phenotypic features and temporal symptom evolution, particularly when structured EHR data are sparse. Despite these advances, most existing literature operate in retrospective settings, focusing on cohort identification, phenotyping validation, or post-hoc prediction tasks where diagnostic signals are already established. In contrast, prospective early detection using only pre-diagnostic EHR history remains relatively underexplored. Identifying patients at risk before formal diagnosis requires modeling subtle intra-visit heterogeneity and longitudinal temporal progression, posing unique methodological challenges that current approaches have not fully addressed.

### B. Deep Learning for Longitudinal EHR Modeling

Deep learning has transformed the analysis of electronic health records, with a wide range of architectures applied across diverse clinical tasks, including information extraction from clinical text, representation learning from structured records, and automated phenotyping [19]. A key driver of this progress is the ability of deep architectures to model high-dimensional, sparse, and temporally structured EHR data and enable learning of longitudinal disease progression patterns across visits, capturing complex nonlinear and temporal dependencies that traditional feature-based machine learning approaches struggle to represent [20]. More recently, Transformer-based architectures have been adapted for EHR data, with BERT-like pretraining on clinical event sequences having been explored [21]–[23]. These architectures demonstrated strong performance in predicting common diseases and next-visit outcomes where diagnostic signals are relatively frequent and prominent. Recent studies have also explored multimodal representation learning for electronic health records by jointly modeling structured clinical variables and unstructured clinical text using transformer architectures and contrastive learning objectives [24]. While these approaches focus on aligning multiple EHR modalities, they typically model clinical events as flattened sequences and do not explicitly capture the hierarchical structure of clinical encounters across visits. However, in the context of rare disease detection, individual visits often contain heterogeneous and partially unrelated diagnoses that introduce substantial noise, and rare disease signals are typically weak and distributed across longitudinal encounters, requiring models to aggregate coordinated phenotypic patterns within visits while capturing their long-term accumulation over time [12], [20]. Motivated by this, our hierarchical approach is specifically designed to reduce intra-visit noise while preserving the longitudinal accumulation of weak phenotypic evidence by treating clinical events within a visit as unordered sets and modeling visits themselves as a temporal sequence.

## III. Dataset

### A. Cohort Construction

We constructed a cohort from an OMOP Common Data Model-formatted clinical data warehouse at the University of Tennessee Health Science Center (UTHSC). The repository integrates longitudinal EHR data from UTHSC and its affiliated institutions, including Le Bonheur Children’s Hospital and Regional One Health, and is part of the Biorepository and Integrative Genomics (BIG) Initiative^1^. The BIG Initiative links structured clinical records with biorepository DNA samples and genomic sequencing data to support precision medicine research in diverse populations across Tennessee [25].

All patients with at least one recorded clinical visit between April 2005 and November 2025 were included, and the final cohort consists of 40,223 patients with 708,422 visits. Patient-level demographics reflect a pediatric-weighted population with the median age 15.7 years and the mean age of 20.6 years. The cohort is 51.1% male, 47.8% female, and 1.1% unknown or other. Racial composition is approximately 48.8% Black or African American and 43.0% White, with 7% identifying as others. The dataset consists of six structured OMOP tables: Patient (demographics), Visit Occurrence (visit metadata), Condition Occurrence (ICD-coded diagnoses), Drug Exposure (medications), Measurement (laboratory results), and Procedure Occurrence. Table I summarizes the scale and key statistics of each table. This study was approved by the Institutional Review Board (IRB) of University of Tennessee with a waiver of informed consent for secondary analysis of de-identified EHR data.

**TABLE I.** Key Statistics of Constructed Cohort by OMOP Table.

| Table | Key Statistics | Value |
| --- | --- | --- |
| Patient | Total patients | 40,223 |
| Visit | Total / Median per patient | 708,422 / 7 |
| Condition | Total / Unique concepts | 2,827,259 / 10,333 |
| Measurement | Total / Numerical fraction | 54,055,276 / 81% |
| Drug | Total / Mean per visit | 6,165,970 / 15 |
| Procedure | Total / Unique procedures | 1,463,170 / 8,935 |
| Overall missingness rate |  | <1.5% (all) |

### B. Data Preprocessing

All condition, drug exposure, measurement, and procedure records were linked to the Visit Occurrence table using patient and visit identifiers within the OMOP CDM schema. Records lacking a valid visit identifier were excluded. Duplicate events within the same visit were removed to prevent redundant representations. Continuous laboratory measurements were discretized into five quantile-based bins derived from the training set and encoded as ordinal integers. Categorical laboratory results (e.g., positive/negative, reactive/non-reactive) were label-encoded. To control computational complexity while preserving clinical coverage, per-visit event counts were truncated at predefined caps, with laboratory events capped at 64, procedures events capped at 32, and total events per visit capped at 128. These thresholds corresponded to the 99th percentile of the empirical per-visit event distributions in the data, thereby retaining the vast majority of observed clinical information while preventing excessively long sequences that increase computational burden. Finally, patient-level visit sequences were assembled in chronological order to construct longitudinal trajectories for modeling.

### C. Rare Disease Phenotyping

Rare diseases are commonly defined as conditions affecting fewer than 1 in 2,000 individuals in the general population according to the European Organization for Rare Diseases (EURORDIS) [26]. In this study, we identified rare disease cases within the EHR using rule-based phenotyping applied to curated condition description strings corresponding to known genetic, congenital, and neurodevelopmental disorders including inborn errors of metabolism, chromosomal abnormalities, neurogenetic disorders, severe congenital malformations, and related phenotypes. Common, non-severe conditions that share terminology with rare genetic diseases (e.g., attention deficit hyperactivity disorder (ADHD), migraine) were explicitly excluded. This approach approximates rare disease identification in structured EHR data where standardized coding for rare conditions is often incomplete or inconsistent. Finally, a total of 3,032 patients were identified as rare disease cases, representing 7.53% of the total population in the cohort.

### D. Prospective Multi-horizon Labeling

For each rare disease case, we identified the date of first recorded diagnosis corresponding to the target rare condition. For each prediction horizon *h* ∈ {7, 30, 90, 180, 365} days, we defined an index date as:

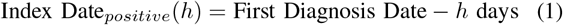

At a given horizon, a patient is labeled positive if a confirmed rare disease diagnosis occurs at least *h* days after the index date. Only EHR data observed strictly prior to the index date are used as model input (Shown as Figure 1). This prospective formulation mirrors real-world deployment: at prediction time, the model has access solely to historical clinical information and no future diagnostic codes, thereby preventing temporal leakage. For those patients without a rare disease diagnosis, the index date is defined relative to their last recorded visit:

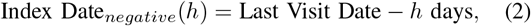

**Fig. 1.**
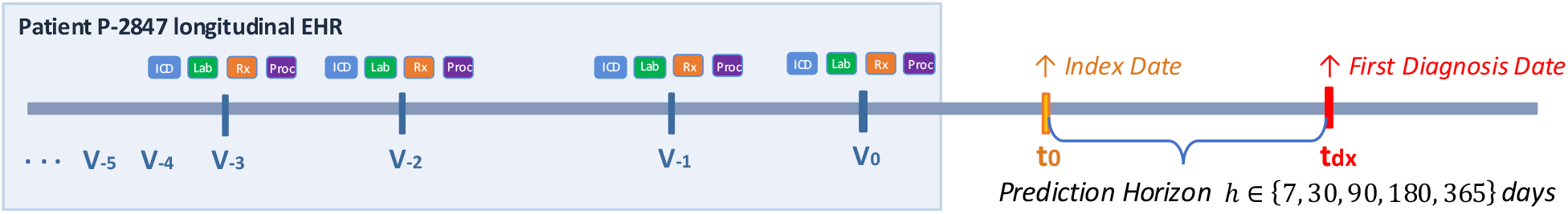
Prospective multi-horizon labeling strategy. For each prediction horizon *h*, the index date *t*_0_ is defined as the first diagnosis date *t*_dx_ −*h*. Only EHR data observed strictly before the index date *t*_0_ are used as model input to prevent temporal leakage, which are clinical events recorded in …, *v*_−5_, *v*_−4_, *v*_−3_, *v*_−2_, *v*_−1_, *v*_0_, and *v*_0_ represents the most recent visit before the index date *t*_0_. Any events recorded after the index date are excluded.

This ensures that each negative patient has at least *h* days of observable follow-up after the index date. Among the 40,223 patients in the cohort, we included only those with at least one clinical visit on or before the horizon-specific index date, ensuring that each patient has non-empty pre-index clinical history available for model input. Therefore, as the prediction horizon increases, both the number of eligible patients and the positive prevalence decrease (Shown in Table II). This also reflects the increasing difficulty of identifying cases further from the time of clinical confirmation, when disease-related phenotypic patterns are less concentrated and more weakly expressed.

**TABLE II.** Final Datasets Characteristics Across Prospective horizons.

| Horizon (days) | Total Eligible | Total Positive | Prevalence (%) |
| --- | --- | --- | --- |
| 7 | 34,106 | 1,762 | 5.17 |
| 30 | 32,520 | 1,653 | 5.08 |
| 90 | 30,067 | 1,439 | 4.79 |
| 180 | 27,799 | 1,252 | 4.50 |
| 365 | 24,425 | 971 | 3.98 |

Note: The count of eligible patients decreases with increasing horizon due to the non-empty pre-index clinical visit requirement.

## IV. Methodology

In this study, we formulated rare disease early detection as patient-level binary classification. Given a patient’s longitudinal EHR data observed up to an index date, the model predicts whether the patient will receive a rare disease diagnosis within the subsequent prediction horizon. In this section, we describe the proposed Hierarchical Set-to-Sequence (HSS) architecture. The whole pipeline is illustrated in Figure 2.

**Fig. 2.**
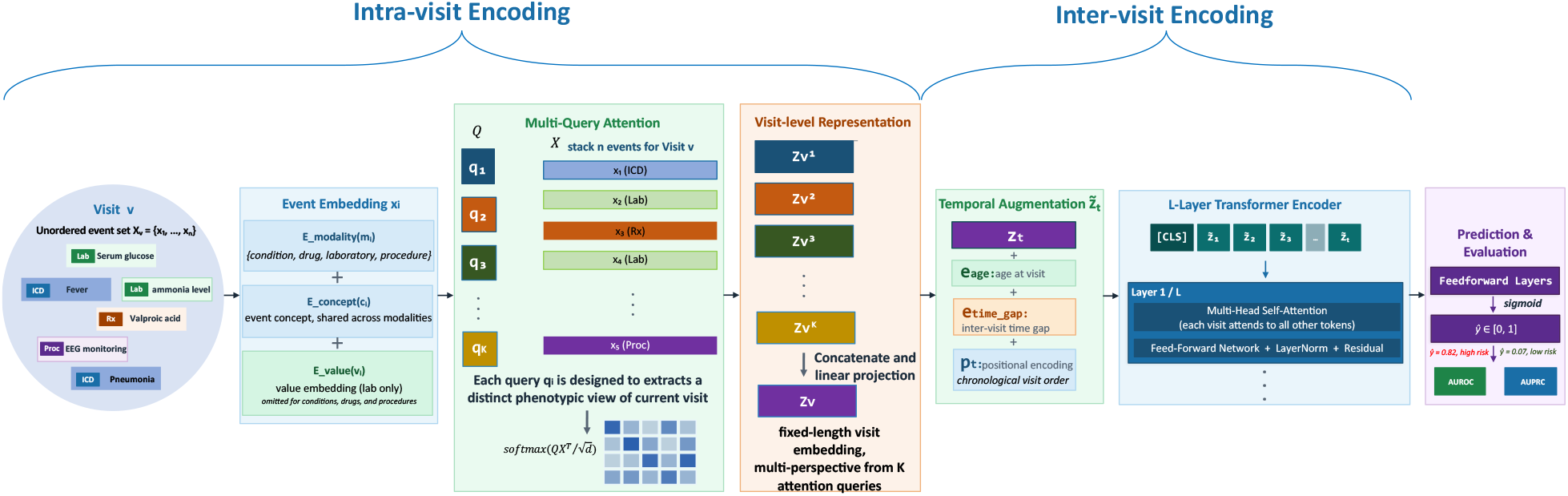
Overview of the proposed Hierarchical Set-to-Sequence (HSS) framework for prospective rare disease detection. Within each clinical visit, heterogeneous events (conditions, procedures, medications, and measurements) are treated as an unordered set and encoded using multi-query attention with *K* learnable phenotype queries to produce a structured visit representation. The resulting sequence of visit embeddings is ordered chronologically and processed by a Transformer encoder to model longitudinal disease progression. The final patient-level representation is used to predict rare disease within the prediction horizon.

### A. Intra-Visit Encoding with Multi-Query Attention

In the context of rare disease trajectory modeling, long-term longitudinal progression across visits is likely to be more informative than the fine-grained ordering of events within a single encounter. Therefore, within each clinical visit, events are modeled as an unordered set of heterogeneous clinical events, including diagnoses, drug exposures, laboratory measurements, and procedures. Let a visit *v* contain *n* events *X*_*v*_ = {*x*_1_, …, *x*_*n*_}. Each event is represented as a composite embedding that captures its clinical concept, modality type, and event value. For an event *x*_*i*_, the embedding vector **x**_*i*_ is constructed as:

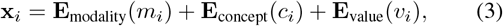

where **E**_modality_(*m*_*i*_) a learned modality embedding that encodes event modality information (i.e., condition, drug, laboratory, or procedure), **E**_concept_(*c*_*i*_) is a learned concept embedding table; all clinical concepts, regardless of their modalities, share a single embedding vector space, allowing cross-modality semantic alignment within a unified latent space. For non-laboratory events (conditions, drugs, procedures), the value embedding **E**_value_(*v*_*i*_), term is omitted, and the event representation consists solely of concept and modality embeddings. Continuous laboratory values are discretized into quantile-based bins and embedded as **E**_value_(*i*), and categorical laboratory values (e.g., positive/negative) are treated as discrete value tokens.

Although intra-visit events are treated as an unordered set, they do not contribute equally to the visit representation because some events are more clinically meaningful than others. Therefore, we adopt multi-head attention pooling with *K* learned query vectors *Q* = {**q**_1_, …, **q**_*K*_} and the attention weight matrix for visit *v* is calculated as:

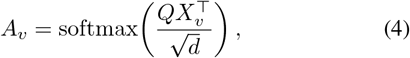

This formulation is inspired by Multi-Query Attention (MQA) [27] but unlike its original auto-regressive decoding mechanism, MQA here serves as a multi-query pooling operator that extracts complementary phenotypic perspectives from heterogeneous clinical events. Each query independently attends over all events to extract a distinct phenotypic representation 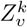 for the current visit:

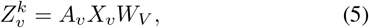

where *W*_*V*_ is a learned value projection. Finally, all *K* query outputs are concatenated and linearly projected to produce a fixed-dimensional visit embedding:

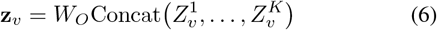

where *W*_*O*_ is a learned projection matrix that maps the concatenated representations to a fixed-dimensional visit embedding. Each query learns to attend to a different subset of events within the visit, allowing the model to simultaneously extract multiple phenotypic perspectives from the same clinical encounter. Importantly, the queries are learned end-to-end rather than predefined clinical categories (e.g., the diagnosis code of each visit). Because rare disease phenotypes are heterogeneous and often span multiple domains, learnable queries allow the model to adaptively discover task-relevant events from data rather than be constrained to predefined clinical categories that may reflect common disease categories and not the underlying rare disease trajectory.

### B. Inter-Visit Temporal Modeling

Given the chronologically ordered sequence of visit representations (**z**_1_, **z**_2_, …, **z**_*T*_ ), we model longitudinal disease progression using a multi-layer Transformer encoder. This design enables flexible modeling of long-range temporal dependencies across clinical visits.

Each visit representation is augmented with two normalized continuous temporal features:

1. Age at visit, encoding disease onset timing since birth.
2. Inter-visit time gap, defined as the elapsed time since the prior visit, capturing irregular temporal dependencies between consecutive clinical visits.

These temporal features allow the model to distinguish between phenotypic trajectories unfolding over months versus years, and to learn that abrupt changes in visit frequency may themselves carry diagnostic signal. Both temporal variables are projected into the visit embedding space through learned linear layers and added to the visit representation:

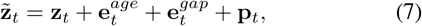

where 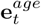 and 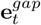 denote projected age at visit and inter-visit time gap embeddings, and **p**_*t*_ denotes sinusoidal positional encoding, representing the chronological order of clinical visits.

The resulting sequence 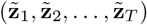 is processed by a *L*-layer Transformer encoder. At each layer, multi-head self-attention enables each visit to attend to all other visits in the patient’s history. This mechanism allows modeling of sparse and temporally dispersed phenotypic patterns that may jointly signal progression toward rare diseases. Finally, the prepended classification token (`[CLS]`) is used as the aggregated longitudinal representation to estimate the probability of disease onset within the prediction horizon.

## V. Results

### A. Experimental Setup

Patients were randomly partitioned at the patient level into training (70%), validation (15%), and test (15%) sets, stratified by the rare disease label to preserve prevalence rate across the splits sets. Model development, hyperparameter tuning, and early stopping were conducted exclusively on the training and validation sets and final performance was evaluated once on the test set. Given the low prevalence of rare diseases across the five prediction-horizon datasets (Shown in Table II), we employed inverse-frequency weighted binary cross-entropy loss [28], which increases the penalty for errors on rare positive patients and reduces bias toward the majority class.

The evaluation metrics included Area Under the Receiver Operating Characteristic Curve (AUROC) and Area Under the Precision-Recall Curve (AUPRC). In rare disease detection where rare disease prevalence is low, AUROC may appear high because patients without rare diseases dominate the population. In contrast, AUPRC focuses on performance among rare disease cases and directly reflects the tradeoff between precision and recall, which is clinically more important when identifying patients for further genetic evaluation. Considering the low prevalence rate of rare diseases in our datasets, our experiments started with a modest number of query vectors (*K* = 8, randomly initialized) and a shallow Transformer depth (*L* = 2), with dropout, weight decay, and early stopping based on the validation AUPRC.

### B. Baselines

We compared the proposed HSS framework against four representative baselines: (1) Logistic Regression, a linear classifier trained on aggregated bag-of-codes features, serving as a strong and interpretable linear baseline, (2) XGBoost, a gradient-boosted decision tree model trained on the same flattened feature representation, capturing nonlinear feature interactions without temporal modeling, (3) BEHRT [21], a transformer-based sequential model that treats diagnosis codes as tokens and models longitudinal disease trajectories using self-attention over flattened sequences of clinical events ordered by visit time, and (4) TransEHR [22], a transformer architecture designed for clinical time-series modeling that embeds structured event triplets (e.g., concept, value, and timestamp) and models temporal dependencies across heterogeneous clinical events. Unlike these transformer baselines which operate on flattened event-level sequences, the proposed HSS framework explicitly models the hierarchical structure of EHR data by first aggregating heterogeneous events within each visit into a unified visit-level representation and then modeling longitudinal dependencies across visits. Together, these baselines enable comparison against linear, tree-based, and the state-of-the-art sequence-based deep learning models for EHR data.

### C. Overall Performance Across Prediction Horizons

Table III compares model performance across prediction horizons for linear, tree-based, sequential, and hierarchical approaches. Among all the baseline models, XGBoost consistently outperforms logistic regression, indicating the presence of non-linear interactions in aggregated clinical features. Transformer-based models (BEHRT and TransEHR) further improve performance over flat baselines, with BEHRT demonstrating slightly stronger performance than TransEHR, suggesting that longitudinal sequence modeling provides additional benefit.

**TABLE III.** Model Performance Comparison Across Prediction Horizons.

| Horizon | Logistic Regression |  | XGBoost |  | BEHRT [21] |  | TransEHR [22] |  | HSS (this paper) |  | Prev. |
| --- | --- | --- | --- | --- | --- | --- | --- | --- | --- | --- | --- |
|  | AUC | PRC | AUC | PRC | AUC | PRC | AUC | PRC | AUC | PRC |  |
| 7 days | 0.845 | 0.359 | 0.852 | 0.407 | 0.891 | 0.557 | 0.889 | 0.534 | **0.893** | **0.601** | 5.17% |
| 30 days | 0.840 | 0.366 | 0.849 | 0.379 | 0.885 | 0.503 | 0.880 | 0.489 | **0.887** | **0.576** | 5.08% |
| 90 days | 0.822 | 0.270 | 0.835 | 0.311 | 0.860 | 0.417 | 0.855 | 0.410 | **0.873** | **0.445** | 4.79% |
| 180 days | 0.807 | 0.247 | 0.810 | 0.261 | 0.831 | 0.316 | 0.812 | 0.308 | **0.858** | **0.331** | 4.50% |
| 365 days | 0.778 | 0.169 | 0.778 | 0.171 | 0.799 | 0.194 | 0.790 | 0.174 | **0.829** | **0.228** | 3.98% |

AUC = area under the receiver operating characteristic curve; PRC = area under the precision-recall curve; Prev. = positive class prevalence at each horizon.

wOur proposed HSS approach achieves the highest AUROC and AUPRC scores at all horizons. Improvements are modest at the 7-day horizon compared to other two transformer baselines (AUPRC 0.601 vs. 0.557 and 0.534); however, at the clinically most challenging 365-day horizon (prevalence 3.98%), HSS achieves AUROC of 0.829 and AUPRC of 0.228, representing improvements of +0.030 AUROC and +0.034 AUPRC over BEHRT, and substantially larger gains over logistic regression and XGBoost baselines. In addition, although the performance of all models degrade with increasing the prediction horizon, the degradation is smaller for our proposed HSS approach compared to all other baseline models.

### D. Additional Analyses

We performed additional analyses to quantify the contribution of the two hierarchical components in the proposed HSS approach. Using the default configuration (*K* = 8 query vectors, *L* = 2 Transformer layers), we performed each of the following change to the HSS architecture and kept everything else identical, specifically:

1. replaced multi-query attention pooling with mean pooling (simple mean of event embeddings per visit) to test whether the learned multi-query attention provides benefit beyond uniform averaging;
2. removed the age-at-visit and inter-visit temporal features during inter-visit encoding to test whether modeling irregular visit gaps provides additional benefit beyond the visit ordering alone;
3. varied the number of query (*K* ∈ {4, 8, 16, 32}) and Transformer depth (*L* ∈ {1, 2, 4, 8} ) to test how the complexity of model structure influences the model performance.

The analyses were conducted at the 365-day horizon, where early phenotypic signals are weakest and hierarchical modeling provides the greatest benefit. Table IV shows the results of additional analyses. As shown in the table, replacing multiquery attention (MQA) with simple mean pooling leads the largest performance drop (ΔAUC = -0.014, ΔPRC = -0.028), suggesting that the structured intra-visit aggregation with MQA provides substantial benefit compared to uniform averaging.

**TABLE IV.**
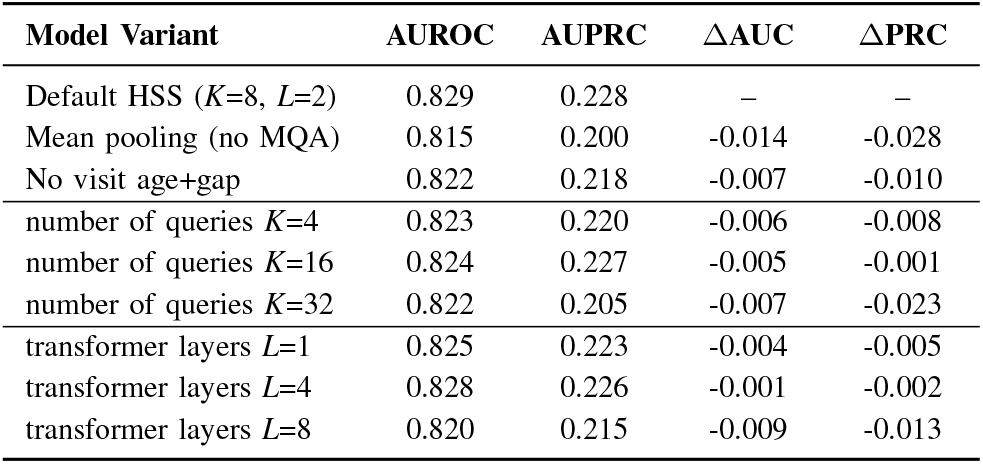
Additional Analyses at 365 Days Prediction Horizon.

Additionally, removing temporal features during inter-visit encoding stage (age-at-visit and inter-visit gap) also reduces model performance, indicating that irregular visit spacing contributes additional predictive information beyond the visit ordering alone. For the optimal number of query vectors (*K*) in MQA and the number of transformer encoder depth *L*, we can observe that the model performance improves from *K* = 4 to *K* = 8, while plateaus at *K* = 16, and degrades at *K* = 32. This suggests that adding more query vectors does not always yield additional performance gain and may instead introduce unnecessary model complexity relative to the limited number of positive cases. Similarly, increasing Transformer depth beyond *L* = 2 does not yield additional gains and leads to performance degradation at *L* = 8, suggesting that shallow temporal modeling is sufficient given limited positive cases and weak long-horizon signals.

## VI. Discussion

In this study, we propose a hierarchical set-to-sequence (HSS) framework specifically designed for prospective rare disease detection using structured EHRs. Because for rare disease trajectory modeling, longitudinal progression across visits is typically more informative than the fine grained ordering of heterogeneous events within a single visit, HSS models intra-visit events as unordered sets and then captures cross visit temporal dependencies through sequence modeling of visit embeddings.

Experimental results show that the proposed HSS approach achieves the strongest performance across all prediction horizons, consistently outperforming linear, tree-based, and Transformer-based baselines in both AUROC and AUPRC. In particular, HSS outperformed BEHRT [21] and TransEHR [22], two state-of-the-art transformer-based frameworks for EHR modeling that operate on flattened token sequences of clinical events instead of separating intra-visit aggregation from inter-visit temporal modeling. These findings suggest that hierarchical modeling is particularly well suited for rare disease detection, in which rare disease detection depends more heavily on structured accumulation of phenotypic evidence across longitudinal visits, where hierarchical modeling appears to be more effective. Importantly, at the 365 days prediction horizon, the performance gap between HSS and baseline models is the largest, which indicates that the relative benefit of hierarchical modeling in our proposed approach increases for longer-horizon rare disease prediction.

The additional analyses further clarify the contribution of each hierarchical component. Replacing multi-query attention with simple mean pooling leads to the largest performance drop compared to the default setting (ΔPRC of -0.028), indicating that structured intra-visit aggregation plays a central role in preserving heterogeneous phenotypic signals. Removing temporal features of age at visit and inter-visit time gap leads to a smaller but consistent performance degradation, suggesting that modeling irregular visit timing provides limited benefit. Increasing the number of queries (*K*) or Transformer depth (*L*) beyond moderate settings does not improve performance. These findings indicate that, in low-prevalence rare disease settings when positive cases are limited, carefully designed hierarchical structure may be more valuable than increasing model complexity.

From a clinical perspective, improved long-horizon performance is particularly relevant for timely referral and diagnostic evaluation. At the 365-day horizon with 3.98% prevalence, an AUPRC of 0.228 represents approximately five times over baseline population risk. This suggests that the model is capable of identifying a subset of patients whose risk is meaningfully higher than the population rate, even one year prior to formal diagnosis. In practice, this level of risk stratification could be clinically meaningful to support prioritized referral for genetic testing and evaluation. Given that many rare disease patients experience prolonged diagnostic delays, even modest improvements in early risk identification may help support more timely evaluation and reduce avoidable diagnostic delays.

This study has several important limitations. First, this study was conducted within a single-institution cohort, and external validation across health systems with different demographic compositions, coding practices, and rare disease prevalence is necessary. Although our cohort includes substantial representation of Black or African American patients, which improves diversity relative to many published EHR studies, differences in diagnostic practice and healthcare access may affect phenotyping accuracy and model generalizability. Future work will investigate integrating the proposed HSS framework into a federated learning setting [29] to enable multi-site validation and improve the robustnewwss and generalizability of rare disease detection models. Second, the positive class in this study was defined using rule based phenotyping rather than genetic variant level confirmation. This inevitably introduced label noise as some patients with undiagnosed disease may remain in the negative class. Prior work has highlighted that EHR-derived labels are often noisy or incomplete, particularly in rare disease settings where diagnostic pathways are complex and heterogeneous, and importantly, handling label noise itself represents an open and challenging research problem [30]. Future studies incorporating whole-exome or genome sequencing data for variant-level validation would provide a more rigorous ground truth and enable more precise performance assessment. Finally, the current analysis relies solely on structured EHR data, while important phenotypic information in rare disease is also often documented in unstructured clinical notes and imaging reports. Recent studies have shown that extracting detailed phenotypic features from clinical narratives can substantially improve rare disease detection performance [6]. Future studies will extend the HSS framework to integrate multimodal data through natural language processing and medical image analysis.

## VII. Conclusion

Early detection of rare diseases from clinical data remains a challenging problem due to the heterogeneous, sparse, and gradually accumulating nature of phenotypic signals across longitudinal visits. In this paper, we introduced the Hierarchical Set-to-Sequence (HSS) framework, designed to reduce intra-visit noise while preserving the longitudinal accumulation of weak phenotypic evidence by treating clinical events within a visit as unordered sets and modeling visits as a temporal sequence. Evaluated on a real-world cohort of 40,223 patients across five prospective prediction horizons ranging from 7 to 365 days, HSS consistently outperforms linear, tree-based, and Transformer-based baselines in both AUROC and AUPRC. Notably, the performance advantage of HSS increases as the prediction horizon extends, with the largest gains observed at 365 days, indicating that the relative benefit of hierarchical modeling increases for longer-horizon rare disease prediction tasks. Future work will extend this framework to multimodal data and external cohorts to further advance prospective rare disease detection.

## Data Availability

All data produced in the present study are available upon reasonable request to the authors

https://uthsc.edu/cbmi/big/

## Acknowledgment

The authors would like to thank Center for Biomedical Informatics at University of Tennessee Health Science Center for their support in cohort construction, data extraction, and regulatory oversight.

## Footnotes

1 https://uthsc.edu/cbmi/big/

